# High-Frequency vs. Low-Frequency Music Therapy in Psychiatric Inpatients: A Randomized Controlled Trial

**DOI:** 10.1101/2025.01.08.25320238

**Authors:** Manuel Esteban-Cárdenas, Ana Gómez-Puentes, Carlos Torres-Delgado, Adrián Hidalgo-Valbuena, Eugenio Ferro

## Abstract

**Introduction:** Music therapy is an evidence-based clinical intervention with demonstrated efficacy in reducing anxiety and depression and in neuromodulation that promotes well-being. Sessions are usually delivered weekly. Psychiatric hospital stays are progressively shorter, requiring interventions to be delivered in brief intensive schemes. Our objective was to evaluate the effectiveness of a high-frequency intervention music therapy (5 sessions/week) compared to a low-frequency (1 session/week) control group.

**Materials and methods:** This is a randomized controlled clinical trial in patients with mental disorders under in-hospital psychiatric treatment. A computer-based random generator created a random allocation sequence to guarantee equal group assignment probability. The primary outcome was symptoms in the Depression/Anxiety/Stress Scale of 21 items (DAS-21). Secondary outcomes were the impact of music therapy sessions on adult patients measured with the CISMA questionnaire (CISMA by Spanish acronyms), and global life satisfaction with the single item for measuring overall life satisfaction (SWLS). To mitigate risks associated with the interventions, at least one healthcare professional will attend each session, alongside a certified music therapist.

The Shapiro-Wilk test was used to test the distribution of variables. We used the Mann-Whitney U test to prove differences between groups and the Wilcoxon signed-rank test differences within groups. Spearman’s test was performed to determine the correlation between continuous variables. Statistical analysis was performed with Jamovi (version 2.6, 2024).

**Results:** Patients in the intervention group had a significant stress level reduction compared to the control group (p=0.033). Both groups experienced significant improvements in anxiety, depression, and global life satisfaction. In addition, positive expectations towards music therapy were associated with greater symptom reduction (r=-0.33, p=0.004).

**Discussion:** To the best of our knowledge, this study is the first randomized controlled trial comparing high-frequency and low-frequency music therapy for psychiatric inpatients. Although both groups demonstrated significant improvement in all the outcomes measured, only the high-frequency group demonstrated greater benefit for stress symptoms. Changes in physiological stress have been reported previously, concordant with our psychological findings. Music therapy had a probable suggestive effect on symptom reduction. These findings highlight the potential of music therapy for stress management in psychiatric hospitals but emphasize the need for further research to standardize session intensity and treatment duration.

**Trial Registration:** ISRCTN registry: ISRCTN87861817 (https://www.isrctn.com/ISRCTN87861817)

## Introduction

The relationship between music and health has been widely studied (1). It has evolved as a complementary intervention for the treatment of mental illnesses and disorders. The American Music Therapy Association (AMTA) defines music therapy as the clinical and evidence-based use of musical interventions to achieve specific therapeutic goals in the treatment of various physical and psychological disorders (2). They are based on the premise that music influences emotional, cognitive, and physical aspects of human well-being, stimulating areas of the brain linked to emotions, memory, and behavior (2).

Music therapy, in addition to demonstrating benefits for people’s general well-being, is used as a complementary therapeutic intervention in the treatment of various physical and mental health conditions (3,4). Its versatility allows it to be applied in different settings, populations, and pathologies (5–7).

It has been demonstrated that music therapy produces modulation of neurochemical systems, promoting relaxation and well-being (6). In addition, it promotes neuroplasticity and the release of neurotransmitters such as dopamine and serotonin, which are related to pleasure, motivation, and well-being (8). Both passive and active participation in its creation have shown positive effects on cognitive and psychosocial functioning (5) as well as reducing pain and anxiety (9).

Previous studies support music therapy as an effective non-pharmacological intervention for emotional regulation, significantly reducing symptoms of anxiety and depression in different populations, including patients with chronic diseases such as those on hemodialysis (10), people with dementia (11) and psychiatric patients, highlighting its impact on improving mood and self-expression (12). It has also been reported that the combination with Progressive Muscle Relaxation (PMR) is effective in reducing anxiety, depression, and stress in patients with breast cancer and gynecological cancer during chemotherapy, also improving the life satisfaction of these patients (13).

However, although music therapy has demonstrated its effectiveness, important questions remain regarding the optimal frequency and duration of the intervention, especially in patients with mental disorders under hospital treatment.

Most music therapy intervention studies with clinical populations contemplate sessions with a weekly frequency and over a relatively long period of several weeks to months. However, psychiatric hospitalizations are currently much shorter, lasting about 10 to 12 days. For this reason, it is necessary to test music therapy interventions that can be administered for brief periods, to be adapted as a complementary treatment to psychiatric hospitalization. To the best of our knowledge, there is no evidence of the benefit of music therapy sessions more frequently than once a week in a hospital setting for mentally ill patients.

The present study aimed to evaluate the effects of high-frequency music therapy (five sessions per week) compared to low-frequency (one session per week) for psychiatric inpatients. We hypothesize that patients receiving a higher frequency of sessions might have better effects on symptoms of anxiety, depression, stress, and global life satisfaction.

## Materials and methods

### Study design

This study is a randomized controlled clinical trial with a parallel group design (the protocol is available at the ISRCTN registry under registration number: ISRCTN87861817). Participants were randomly assigned to either the high-frequency music therapy intervention group or the low-frequency control group. The high-frequency intervention group received five sessions of music therapy in one week of inpatient treatment at a rate of one session per day for five consecutive days. The low-frequency control group received one music therapy session in one week of inpatient treatment.

The primary outcome was symptoms of depression, anxiety, and stress during inpatient treatment with the Depression/Anxiety/Stress Scale of 21 items (DAS-21). Secondary outcomes were the impact of music therapy sessions on adult patients measured with the CISMA questionnaire (CISMA by Spanish acronyms), and global life satisfaction.

### Participants

The sample size was calculated with a power of 85% and a bilateral significance level of 0.05, aiming to detect a mean difference of 4 between the experimental and control groups, with an estimated standard deviation of 5. A minimum sample size of 60 participants (30 per group) was targeted, with 35 per group to account for potential dropouts. Participants were receiving inpatient treatment in a tertiary referral medical center for psychiatry in Bogotá, Colombia (ICSN – Clínica Montserrat – Hospital Universitario) during their participation in the study.

Inclusion criteria were: 1) individuals over 18 years of age; 2) more than 48 hours of psychiatric hospitalization at the time of recruitment; 3) pharmacological treatment with medication adjustments in the last two weeks. Exclusion criteria were: 1) previous participation in music therapy programs; 2) main diagnosis of hospitalization abstinence syndrome or substance dependence; and 3) more than six days of hospitalization at the time of selection.

The recruitment period for this study started on August 1, 2024, and ended on September 1, 2024, because the sample was completed. The interventions, including follow-ups, were completed by September 9, 2024. During the recruitment period, 106 patients undergoing inpatient treatment were identified. 91 participants signed written informed consent forms and were randomly assigned to either the high-frequency intervention group or the low-frequency music therapy control group (Figure 1).

**Fig 1.**
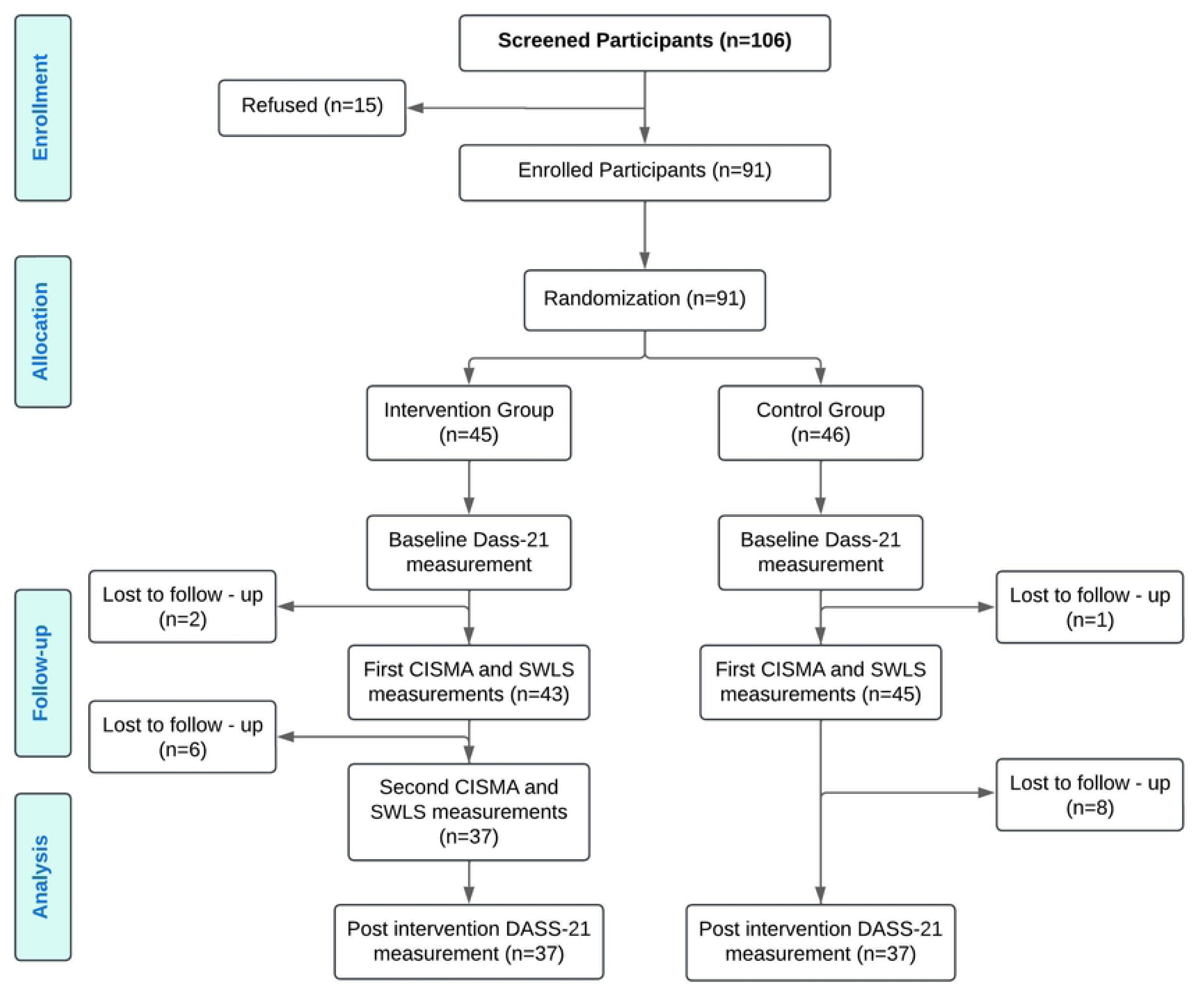
Flow chart of study participation. The number of patients recruited and how many completed each study phase is shown. Source: Own elaboration.

The study began with the recruitment of 106 participants, of whom 15 refused to participate, resulting in a final sample size of 91 enrolled participants. These participants were randomized into two groups: the intervention group (n=45) and the control group (n=46). Baseline measurements using the DASS-21 tool were conducted for both groups. During follow-up, 2 participants in the intervention group and 1 in the control group were lost before the first CISMA and SWLS measurements, leaving 43 and 45 participants, respectively. In the second follow-up an additional 6 participants lost in the intervention group and 8 in the control group, resulting in 37 participants remaining in each group for the second CISMA and SWLS measurements and post-intervention DASS-21 assessment.

### Measuring instruments

#### Depression, Anxiety, and Stress Scale - 21 (DASS-21)

Evaluates the levels of depression, anxiety, and stress-associated symptoms, with scores categorized according to symptom severity (14,15). This self-applied scale has been validated in various populations and cultures, including the Colombian population (15,16). It has been found to have adequate psychometric properties, such as high internal consistency and a stable factor structure (Cronbach’s alpha 0.94 for the depression subscale, 0.87 for the anxiety subscale, and 0.91 for the stress subscale) (14).

#### Questionnaire of the Impact of Music Therapy Sessions in Adults (CISMA)

An innovative self-report tool, designed by music therapists to assess general well-being before and after the music therapy session, evaluating aspects such as mood, physical pain, and relaxation (17). A self-administered questionnaire, simple and short, provides an overview of the impact experienced by the patient in the session and evidences the overall well-being of the individual.

#### Single item for measuring overall life satisfaction (SWLS)

Measures the patient’s general perception of satisfaction with life, using a single item validated in multiple studies (18).

### Procedure

After providing written informed consent, demographic data were collected. Participants were then randomly assigned in a 1:1 ratio to either the high-frequency music therapy intervention group or the low-frequency control group. The random allocation sequence was generated using a computer-based random number generator to ensure an equal probability of group assignment. The study employed simple randomization, ensuring that each participant had an equal probability of being assigned to either group. No restrictions, such as blocking or stratification, were applied during the randomization process.

Both groups took baseline and post-intervention measurements of the DASS-21, life satisfaction (SWLS), and CISMA questionnaire. Participants in the high-frequency intervention group received five music therapy sessions, with a frequency of one session per day for five consecutive days. Participants in the control group received only one music therapy session during one week of hospitalization (Fig 1).

The random allocation sequence was generated by an independent statistician using a computer-based random number generator. Participants were enrolled by a blinded study coordinator who did not have access to the allocation sequence. Group assignments were made by a research assistant not involved in recruitment or intervention delivery.

### Structure of the sessions

The structure of the music therapy sessions had a progressive approach integrating musical interaction with relaxation and emotional synchronization techniques. Each session lasted 30 minutes, organized into four moments: warm-up, group synchronization, main activity, and closing. The use of the PLAYTRON MIDI controller, together with the piano and guitar, allowed stimulation of both the creativity and attention of the participants. In addition, the use of conductive objects such as fruits, water, and plants facilitated multisensory interaction, adding a playful and exploratory dimension to the sessions.

In the first part of the session, we sought mutual recognition among the participants and the establishment of group cohesion. A welcome song was used to help create a space of trust and openness. Repetition of the same song or piece of music in each session allows patients to associate the melody with a safe and familiar environment, facilitating the transition to the “here and now” and the focus on group activity.

Progressive Muscle Relaxation (PMR) was the focus of the second moment, where participants worked on synchronizing their breathing with muscle activation and relaxation. Using thoracic breathing, patients are invited to “scan” their body for tension and then focus on relieving those areas. This exercise prepares participants for creative and musical work while encouraging emotional and physical regulation.

In the main moment of the session, the PLAYTRON, a MIDI controller that allows participants to interact with everyday objects turned into musical instruments, is introduced. This activity encourages sustained attention and role-playing within the group, as well as encouraging creativity and sensory exploration. PLAYTRON helps patients connect with their environment in a novel way, expanding their capacity for wonder and contributing to the regulation of catastrophic thoughts.

To close the session, a farewell song was used, combining the voices of the participants with the sounds created by the PLAYTRON. This musical closure serves as a ritual of completion, where participants say goodbye to the group and the activity, while the multisensory environment, with the aromas of the fruits and plants used, plus the possibility of experiencing the taste of their instrument, accompanies this moment. This final activity reinforces group cohesion and provides a sense of positive emotional closure for the participants.

### Ethics statement

This study was conducted following the ethical guidelines established in the Declaration of Helsinki of the World Medical Association (19) and the Colombian legislation for research involving human participants (20).

Both the research protocol and the informed consent were approved by an independent research ethics committee (CEI Campo Abierto Ltda, approval act No. 209 of July 2024). All study participants signed the informed consent document before any research procedure was performed. In addition, to guarantee the principle of equity of the participants assigned to the control group and considering that they received fewer music therapy sessions than the high-frequency group, once the measurement of study outcomes was completed, they were allowed access to the opposite intensity of sessions.

To ensure nonmaleficence, the investigators who applied for the tests considered immediately suspending the study when they noticed any risk or harm to the health of the participants or discontinuing the study on an individual basis when they noticed a particular risk in a research participant.

### Data analysis

A descriptive analysis characterized the sample, including measures of central tendency (mean, median) and dispersion (percentiles). The normality of the continuous variables was evaluated using the Shapiro-Wilk test. Since all the variables presented a non-normal distribution (p < 0.05), nonparametric tests were used for the statistical analysis, establishing a significance level of p < 0.05 in their initial or final values.

We used the Mann-Whitney U test to prove the differences between the intervention and control group. The Wilcoxon signed-rank test was applied to evaluate differences within groups for pre- and post-intervention measurements. Spearman’s test was performed to determine the correlation between continuous variables. Statistical analysis was performed with Jamovi (version 2.6, 2024).

## Results

A total of 106 patients were screened, of which 91 participants were enrolled and randomly assigned. Figure 1 describes the recruitment and follow-up of study participants. For the primary outcome analysis, 37 participants in the intervention group and 37 in the control group were included. Table 1 presents the demographic and clinical characteristics of the sample. As can be seen, no significant baseline differences were found between the assignment groups regarding gender, schooling, diagnostic axis, and expectation of music therapy (Table 1).

**Table 1.** Demographic and clinical characteristics.

|  |  | **Intervention**<br>**Group**<br>**(n=37)**<br>**n (%)** | **Control Group**<br>**(n=37)**<br>**n (%)** | **P Value\*** |
| --- | --- | --- | --- | --- |
| Age | Mean (S.D) | 37.5 (±37.5) | 34.9 (±16.3) | 0.307 |
| Gender | Male | 13 (35) | 6 (16) | 0.11 |
|  | Female | 24 (65) | 31 (84) |  |
| Level of<br>education | Postgraduate | 15 (20) | 10 (14) | 0.457 |
|  | Professional | 5 (7) | 9 (12) |  |
|  | Bachelor's,<br>Technological | 15 (20) | 17 (23) |  |
|  | Less than<br>Bachelor's | 2 (3) | 1 (1) |  |
| Expectations of<br>music therapy | Very Positive | 21 (5) | 25 (68) | 0.43 |
|  | Positive | 11 (30) | 10 (27) |  |
|  | Neutral | 5 (13) | 2 (5) |  |
| Diagnostic<br>group | Depressive<br>Disorder | 23 (62) | 22 (59) | 0.602 |
|  | Bipolar Disorder | 7 (19) | 9 (24) |  |
|  | Anxiety/Stress<br>Disorder | 2 (5) | 4 (11) |  |
|  | Psychosis | 4 (11) | 1 (3) |  |
|  | Other | 1 (3) | 1 (3) |  |

\*Mann-Whitney test.

Regarding the primary outcome, post-intervention measurements in the high-frequency group showed a greater decrease in the stress subscale of the DASS-21, with a statistically significant difference (p=0.033) and a low effect size (Coheńs d=0.5), compared to the control group. This indicates that the high-frequency intervention was more effective in reducing stress levels (Fig 2). There were no differences in the anxiety and depression domains in the post-intervention measurements (p=0.21 and p=0.34 respectively). There were also no differences between groups in CISMA secondary outcome measures (p=0.32) or global life satisfaction (p=0.31) (Fig 2).

**Fig 2.**
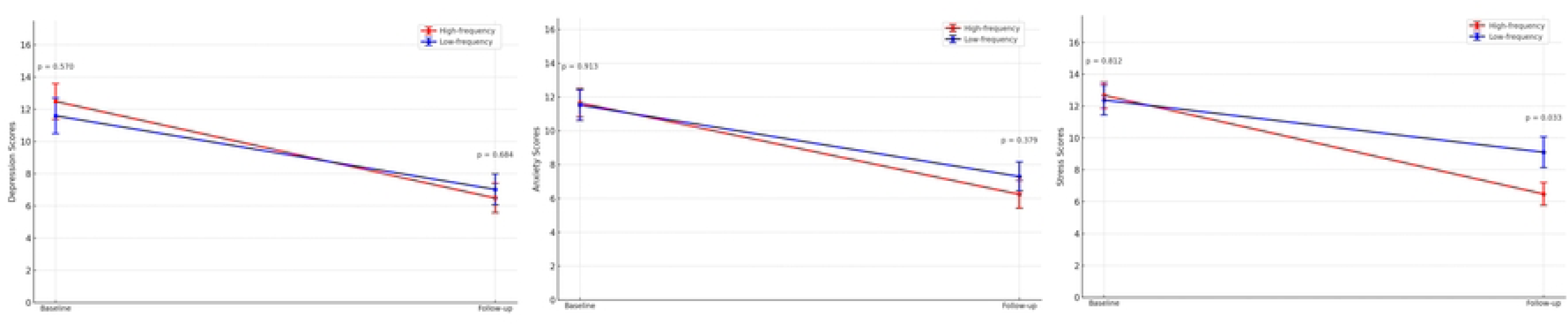
Comparison of DASS-21 between groups. The difference in slopes in response to depression, anxiety, and stress in both groups. Mann-Whitney test.

Regarding the participants’ expectations of music therapy, the majority reported having a very positive expectation (n=46, 62%), followed by a positive expectation (n=21, 28%) and neutral (n=7, 10%). None of the participants reported having a negative or very negative expectation about music therapy, and no significant differences were observed between the high and low-frequency groups in terms of their initial expectations of the intervention (p=0.43).

Positive correlations were found, employing a matrix, between the expectation of music therapy and final life satisfaction (P<0.001), CISMA final total score in the first (P=0.005) and last measurement (P=0.001). Negative correlations were observed between expectancy and symptoms assessed by the Dass-21 except for the anxiety subscale (P=0.093). These results suggest that the expectation held about the music therapy intervention may positively influence its results. No correlations were found between the outcomes measured and variables such as gender, age, schooling, and the diagnostic axis (Table 3).

**Table 2.**
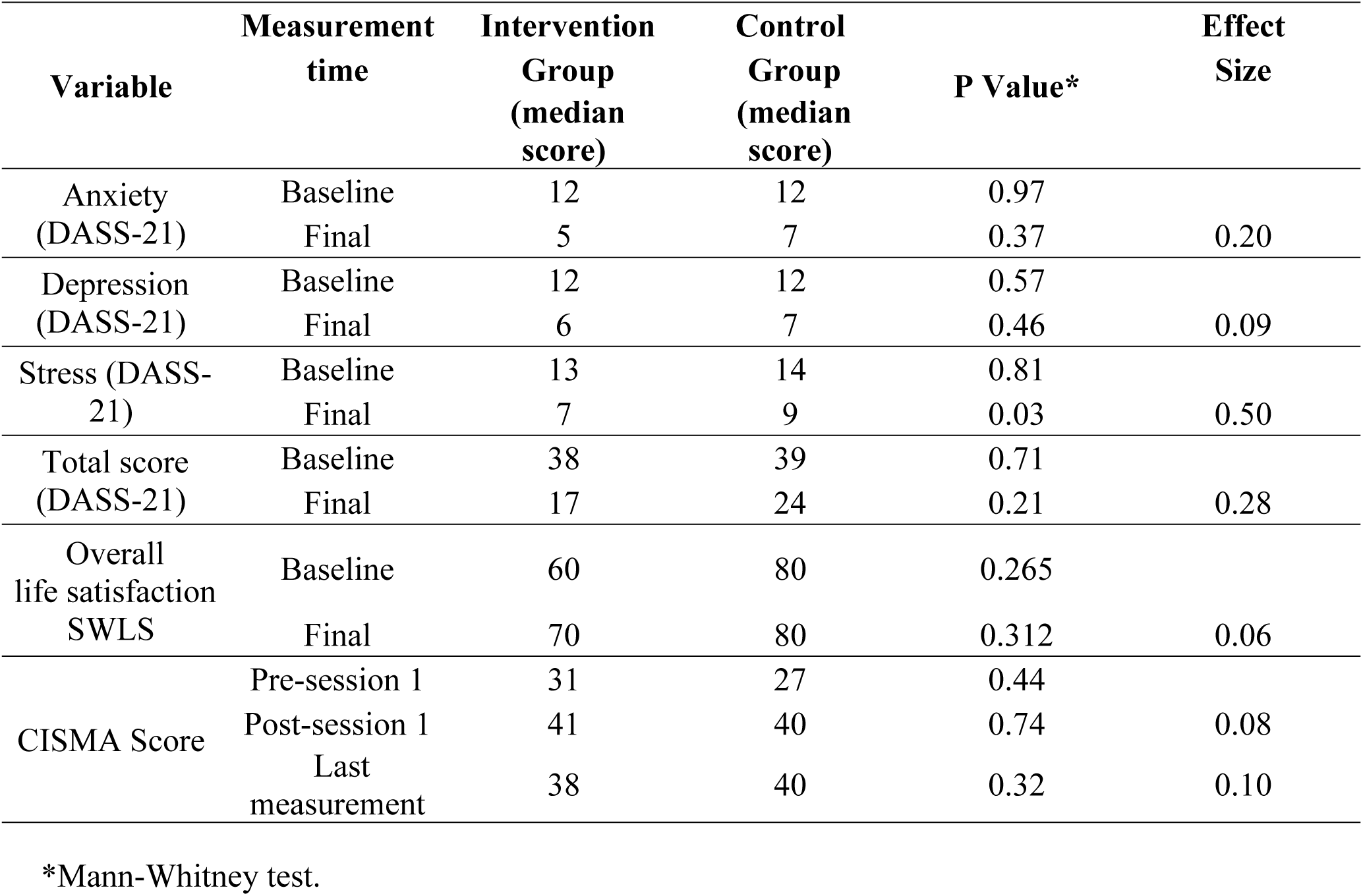
Measurement of outcomes at baseline and follow-up.

**Table 3.** Correlations between levels of expectation and outcomes.

| Outcome | Rho<br>Spearman | P Value |
| --- | --- | --- |
| Final Anxiety (DASS-21) | -0.197 | 0.093 |
| Final Depression (DASS-21) | -0.361 | 0.002 |
| Final Stress (DASS-21) | -0.318 | 0.006 |
| Final DASS-21 Total score | -0.330 | 0.004 |
| Final Global Life Satisfaction | 0.431 | <0.001 |
| CISMA Score Post-Session 1 | 0.323 | 0.005 |
| CISMA Score Post-Session 3 | 0.286 | 0.101 |
| CISMA Score Last<br>Measurement | 0.363 | 0.001 |

Values (P<0.05) show correlations between expectancy and outcome, while positive Spearman’s Rho values suggest a direct correlation and negative values, an inverse one.

Paired sample analyses within each group as well as in the whole sample showed significant differences in the reduction of all subscales of DASS-21 and the total DASS-21 score, and an increase in general life satisfaction as well as feelings associated with music therapy (P<0.001) (Fig. 3).

**Fig 3.**
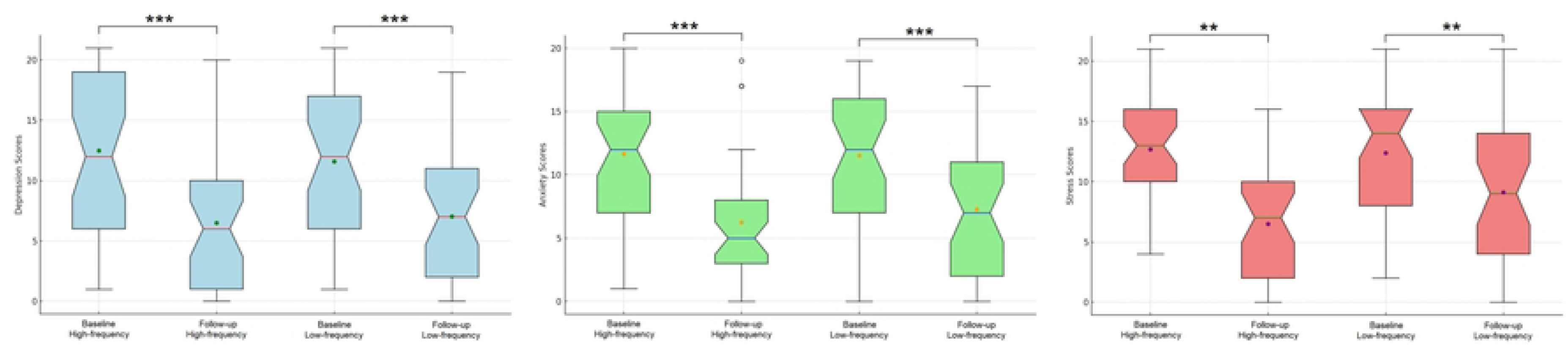
DASS-21 domains intragroup comparisons. Anxiety, depression, and stress scores showed significant decreases both in the high-frequency group and low-frequency group between baseline and follow-up. **p=0.001 ***p<0.001

No adverse events were observed or reported by participants in either the intervention or control group during the study.

## Discussion

To the best of our knowledge, this is the first randomized controlled clinical trial to evaluate the effectiveness of high-frequency music therapy compared to low-frequency music therapy as part of inpatient psychiatric treatment. We found that high-frequency music therapy was more effective in the symptomatic domain of stress than low frequency, with a low effect size. We found no significant differences in symptoms of depression, anxiety, subjective assessments of life satisfaction, or perception of music therapy between groups.

Although there was a significant improvement in all outcomes measured in both groups, there was no group without music therapy intervention, so it cannot be suggested that this improvement is due to music therapy intervention since patients had other treatments as part of psychiatric hospitalization, including psychotropic drugs. Additionally, in this study, we found the influence of a suggestive effect of music therapy, since the participants who reported greater expectations before music therapy sessions had better outcomes in depressive symptoms, stress-associated, global life satisfaction, and perception of the impact of music therapy.

When evaluating the intensity of sessions needed to find an effect, the meta-analysis of Aalbers et al. found that adding twelve music therapy sessions to the usual treatment is more effective in reducing symptoms of depression, anxiety, and improved functioning (21), which supports the results of our study in terms of high-intensity music therapy interventions. Similarly, the study by Lee et al., which applied between one and seven weekly sessions, showed improvements in walking speed and mobility in patients with Parkinson’s disease (22), a range of sessions comparable to that of our study.

There is still a lack of evidence that establishes the minimum intensity of sessions necessary to generate a significant effect with music therapy. Colin et al. found that a daily 30-minute music listening for three weeks reduced stress levels in geriatric healthcare workers (23). This time was similar to the session length of our study but was not structured music therapy sessions, although, finding similar results in stress outcome.

These findings are consistent with previous studies suggesting that music has a positive impact on patient’s emotional well-being, as in the study by Shabandokht-Zarmi et al. that has shown that music has a calming effect and can reduce pain and anxiety in patients during medical procedures (10). As in our study, patients who received both high-frequency and low-frequency intervention reported lower levels of anxiety on post-intervention measurements. These results are aligned with previous research, as noted in the systematic reviews included by Tang et al. and Zhao et al., where there is evidence that the duration and volume of musical treatment significantly influence emotional outcomes (10,24).

The study by Zhou et al. demonstrated how the combination of musical interventions (MI) with Progressive Muscle Relaxation (PMR) is effective in treating psychological problems in particular populations, such as breast and gynecological cancer patients undergoing chemotherapy, where improvements in anxiety, depression and overall quality of life were observed (25). The combination of MI and PMR has been associated with significant reductions in cortisol levels, a physiological marker of stress, and improvements in emotional well-being and quality of life. This music intervention structure was the same that we performed in our study, with concordant results in stress outcomes, but measured in psychological variables of stress.

Evidence indicates that music can reduce stress and anxiety by modulating neuroendocrine responses, acting on the peripheral nervous system and hippocampus, and stimulating the release of neurotransmitters such as dopamine and serotonin, which contribute to mood regulation (26). These biological mechanisms could explain the improvements observed in our study, particularly in patients in the high-frequency group, who showed a greater reduction in stress levels, a trend similar to that observed in previous studies using combined MI and PMR interventions (25,26).

In association with our findings concerning the improvement of stress, although there are no studies on inpatients, a decrease in stress has been recorded in operating room staff associated with music therapy (27) and likewise as an acute response of students before facing an exam (28) and in patients with osteosarcoma (29). Likewise, there was evidence of improvement in anxiety and stress in labor and cesarean section patients (9). These findings strengthen the model proposed by our high-frequency intervention and the results obtained in our study.

Studies have also demonstrated the effectiveness of music therapy in reducing depression and anxiety in various populations, as shown by Jingya Zhang et al. (2023) who in their review of 19 studies, found that musical intervention had a significant effect in reducing depressive and anxious symptoms in people with dementia (PLWD) who present symptoms as a result of neuropathological alterations associated with their disease, suggesting that music therapy in addition to being an accessible and safe method, is also an effective non-pharmacological method for emotional regulation (23). These findings support our study, in that our entire sample presented improvement in all the variables measured, although they were not compared with patients who did not undergo intervention.

The pilot study by AE-NA Choi et. al conducted with a group of hospitalized psychiatric patients, showed a reduction of depressive, anxious, and other psychological symptoms with group music therapy interventions where they point out that music can activate neural networks leading to better physical and psychological function, as well as the combination of sensory stimulation through active participation with instruments and analysis of lyrics, allowed patients to express their emotions and increase their self-esteem, which contributed to improve stress responses, reduce anxiety and improve overall mood (13). These findings strengthen our study, where although the difference in results associated with the intensity of the music therapies is limited to changes in symptoms associated with stress. All of our measured variables improve which strengthens a biological and replicable basis of the response to the intervention.

In our study, no significant differences were observed between the two groups in other outcomes such as global life satisfaction, which coincides with the findings of Lee et al., which also found no significant effects on quality of life after music therapy in geriatric patients (6), as well as that reported by Aalbers et al. who found benefits in depression, anxiety, but not in quality of life in patients with depressive disorder (21). The meta-analysis of Bradt et al. showed reductions in anxiety and depression in breast cancer patients (30), and Tao et al. demonstrated a decrease in anxiety after 8 weeks of intervention in combination with mindfulness (3). On the other hand, doubts arise regarding new studies that can deepen the physiological factors associated with these findings, studies such as Umbrello et al. who showed that a single 20-minute session of music therapy was sufficient to reduce heart rate, respiratory rate and blood pressure in patients on mechanical ventilation (31). This finding is congruent with the study by Han et al., who recorded similar outcomes in mechanically ventilated patients (32).

The patient’s positive expectations towards music therapy seem to have significantly influenced the results, underlining the relevance of psychological factors in the effectiveness of these interventions. This result is consistent with previous studies, such as that of Zhong et al., which demonstrated greater adherence and better outcomes in patients with expectations of improvement after music interventions (20).

Limitations of this trial are that it was conducted in a single-center setting, which may limit generalizability. Our trial was conducted in a single urban psychiatric referral center, which may not be replicated in other clinical settings such as ambulatory treatment, general hospitals or rural areas. Potential sources of bias include the inability to blind participants due to the nature of the intervention, which may introduce performance bias. This fact was even observed in our results in the finding of correlation of the main outcome with the expectancy of music therapy. Therefore, the results should be interpreted with caution, and future controlled studies that address these inherent limitations of the design should be considered.

Furthermore, the use of self-reports and observation by the music therapist may have generated biases of social desirability and subjectivity, affecting the validity of the results. Despite using validated scales such as the DASS-21 and the global life satisfaction scale, the CISMA scale is still under development, limiting the measurement’s robustness. In addition, our sample included the majority of participants with depressive and bipolar disorders, which have to be taken into account for the generalizability of the results. It is suggested that future research should include additional objective measures and a more diverse sample to strengthen the validity and applicability of these findings.

No adverse effects were reported during the study, indicating that the intervention was safe and well-tolerated. These findings suggest a favorable balance between benefits and potential harm, supporting the clinical applicability of the intervention.

## Data Availability

All data files are available from the OSF Storage database (Ferro, E. 2024, October 15). High-Frequency vs Low-Frequency Music Therapy in Psychiatric Inpatients: A Randomized Controlled Trial. https://doi.org/10.17605/OSF.IO/384FB

## Acknowledgments

Primarily to our patients, who gave us their time and opportunity to contribute to the growth of knowledge. To our entire team, made up of our therapist, medical support staff, researchers, and consultants. As well as to the Clinica Montserrat – Hospital Universitario, for offering its disposition and facilities for the planning, execution, and analysis of this study.

## Author Contributions

Conceived and designed the trial: MEC, AGP, E-F, CTD. Random allocation sequence: E-F. Enrolled participants: AGP. Assigned participants to interventions: MEC. Execution of intervention: AHV, MEC, AGP. Data collection: MEC, AGP. Data analysis: E-F, CTD, MEC.

Data interpretation: MEC, AGP, E-F, CTD. Wrote the manuscript: MEC, AGP, E-F, CTD. Critical review and approval of manuscript: MEC, E-F, CTD, AGP, AHV.

## Funding

This study was funded by the Instituto Colombiano del Sistema Nervioso – Clínica Montserrat. The funding agency had no role in the design, collection, analysis, or interpretation of data; report writing; or decision to submit the article for publication. No additional external funding was received for this study.

## Notes

### Competing Interest Statement

The authors have declared no competing interest.

### Clinical Trial

ISRCTN87861817

### Funding Statement

The author(s) received no specific funding for this work.

### Author Declarations

On July 26, 2024, as recorded in Act 209, "The Ethics Committee in Research of C.E.I. CAMPO ABIERTO LTDA" reviewed and approved Research protocol, Informed consent document,and all instruments. C.E.I Campo Abierto LTDA. Tel: +57 310 697 4943

